# A Competing Risk Analysis of Early COVID-19 Treatments

**DOI:** 10.1101/2021.12.16.21267866

**Authors:** Gilberto Gonzalez-Arroyo, Mario F. Rodriguez-Moran, Maricela Garcia-Arreola, Karla Guadalupe Lopez-Lopez, Tonatihu Ortiz-Castillo, Salvador Gomez-Garcia, Cleto Alvarez-Aguilar, Anel Gomez-Garcia, Arturo Lopez-Pineda

**Affiliations:** Amphora Health, Morelia, Michoacan, Mexico; Decentralized Administrative Operation Organ, Mexican Institute for Social Security (IMSS), Morelia, Michoacan, Mexico; Decentralized Administrative Operation Organ, Mexican Institute for Social Security (IMSS), Monterrey, Nuevo Leon, Mexico; Faculty of Medical and Biological Sciences. Universidad Michoacana de San Nicolas de Hidalgo (UMSNH), Morelia, Michoacan, Mexico; Biomedical Research Center of Michoacan, Mexican Institute for Social Security (IMSS), Morelia, Michoacan, Mexico

**Keywords:** COVID-19, Survival Analysis, Machine Learning, Real World Data, Mexico

## Abstract

**Introduction:** The advent of the SARS-CoV-2 virus posed formidable challenges on a global scale. In the year 2020, existing treatments were not tailored specifically to combat this novel virus, and the absence of a developed vaccine added to the complexity. Clinical guidelines underwent rapid evolution during the initial months of the pandemic, leaving uncertainty about the efficacy of various drug combinations in treating the disease. This study delves into an analysis of outcomes during the early stages of the pandemic within the Mexican Institute of Social Security (IMSS), the largest healthcare system in Mexico.

**Material and Methods:** In this retrospective observational study, we examined the medical records of 130,216 COVID-19 patients treated in two Mexican states throughout the year 2020. We conducted a competing risk analysis, considering death and recovery as potential outcomes. This was further complemented by a Cox-regression and Kaplan-Meier analysis. To enhance predictive insights, machine learning models were constructed to forecast outcomes at 10, 20, and 30 days.

**Results:** Our analysis revealed a heightened prevalence of comorbidities, including obesity, diabetes, and heart disease, aligning with Mexico’s established epidemiological profile. Mortality patterns indicated occurrences approximately 15-20 days from the onset of symptoms. Notably, patients undergoing treatment with cephalosporin in conjunction with neuraminidase inhibitors (NAIs) exhibited the poorest survival rates, whereas those receiving adamantane, fluoroquinolone, or penicillin demonstrated the most favorable survival outcomes.

**Conclusions:** The identified associations caution against the utilization of specific treatment combinations, providing crucial insights for refining the country’s clinical guidelines and optimizing patient care strategies.

## 1. Background

Over the past few years, there has been a comprehensive delineation of the clinical and epidemiological profiles of individuals affected by COVID-19 (*1*). Yet, in the initial phase of the pandemic, SARS-CoV-2 infections were primarily identifiable through a range of clinical manifestations, varying from mild to severe cases, and even progressing to potentially fatal pneumonias. This severity often required admission to intensive care units, contributing to elevated global mortality rates (*2*). As of the present date, the enduring consequences of COVID-19 continue to manifest in a myriad of organs and systems, underscoring the long-term impact of the disease (*3*)

Since the emergence of COVID-19, significant strides have been made in refining treatment protocols, reflecting an enhanced understanding of the virus and its pathophysiology. Initially marked by supportive care measures (*4*), treatment approaches have evolved to incorporate antiviral medications, immunomodulators, and novel therapeutic strategies (*5*). Notably, advancements in the use of monoclonal antibodies, antiviral drugs like remdesivir, and the exploration of immunomodulatory agents such as tocilizumab have demonstrated promising outcomes in managing severe cases (*6*). However, in the early days of the pandemic back in March 202, guidelines and available treatments were not well understood. The rapidly evolving nature of the novel coronavirus, coupled with limited knowledge of its behavior and transmission patterns, posed formidable challenges for healthcare professionals and policymakers alike. Treatment strategies were exploratory, and clinical guidelines underwent frequent revisions as the scientific community garnered more insights into the virus.

Mexico faced a significant challenge in the early days of the pandemic, grappling with one of the highest observed case fatality rates globally at 7.6%, notably surpassing the 2.1% global average (*7*). During this critical period, policy and guidelines underwent iterative revisions in response to the dynamic landscape of the pandemic. The initial guideline, published on April 7, 2020, recommended a spectrum of treatments including hydroxychloroquine, azithromycin, steroids, human immunoglobulin, interferon beta b1, IL-6 inhibitors, and JAK inhibitors, with a caveat for clinical evaluation (*8*). Subsequent updates, particularly on June 25 and September 25, 2020, underscored a shift in focus toward evaluating complementary therapies, research protocols, and support for managing comorbidities (*9*, *10*).

Despite early optimism, studies exploring hydroxychloroquine demonstrated no discernible prophylactic effects (*11*). Similarly, in hospitalized COVID-19 patients, whether treated with hydroxychloroquine alone or in combination, no significant improvements or reductions in hospitalization duration were observed (*12*, *13*). Comparable results emerged with the use of convalescent plasma in patients with severe pneumonia secondary to SARS-CoV-2 (*14*). Ongoing experimentation involving neutralizing antibodies (*15*) and the therapeutic potential of B-1A cells (*16*) suggested promising avenues, but their effectiveness and accessibility still necessitate further in-depth scrutiny through additional studies. This dynamic evolution of treatment strategies underscores the complexities inherent in navigating the uncertainties of an evolving pandemic landscape.

As the global community faced the challenges posed by the novel coronavirus, healthcare professionals in Mexico, and many countries, navigated a complex landscape of evolving treatment guidelines and a dynamic understanding of the virus. This competitive risk analysis delves into the outcomes of different treatment strategies, shedding light on their impact on two events, death and recovery. mortality rates, hospitalization durations, and recovery rates. Our analysis aims to extract valuable lessons from Mexico’s pandemic response, providing insights that transcend borders and inform future healthcare strategies amid global health challenges.

## 2. Materials and Methods

### Study design

This was a retrospective electronic cohort study to analyze the effect of real-world treatments in COVID-19 patients seen at the Mexican Institute for Social Security (*Instituto Mexicano del Seguro Social*, IMSS), the largest healthcare provider in Mexico. The patients were seen at IMSS facilities between March 1st, 2020, and March 1st, 2021 in two states, Michoacan (West) and Nuevo Leon (North). IMSS employs a National Biosurveillance System for Infectious Diseases (*Sistema Nacional de Vigilancia Epidemiológica*, SINOLAVE), which collects relevant information about patients, including demographics, initial symptoms, comorbidities, testing procedures and results, initial medication intake, mortality, and discharge.

Our study has two components, first, we aimed to understand the various treatment patterns used to treat COVID-19 patients, evaluating their performance with mortality as the main outcome and recovery as a secondary outcome taking the latter as the situation when the patient is no longer ill or is discharged because their symptoms improved. Secondly, we then build two machine learning (ML) models to predict the mortality or recovery of a patient given the available data at a given time point of disease progression (10, 20, and 30 days after admission).

A diagram of our study design is shown in Figure 1. Our analysis was run using the R programming language with multiple libraries, which are displayed in a public repository (https://github.com/AmphoraHealth/covid19). Institutional Review Board (IRB) approval was obtained by IMSS National Bioethics Committee and IMSS National Research Committee, under protocol numbers R-2021-1912-014, R-2020-785-058.

**Figure 1.**
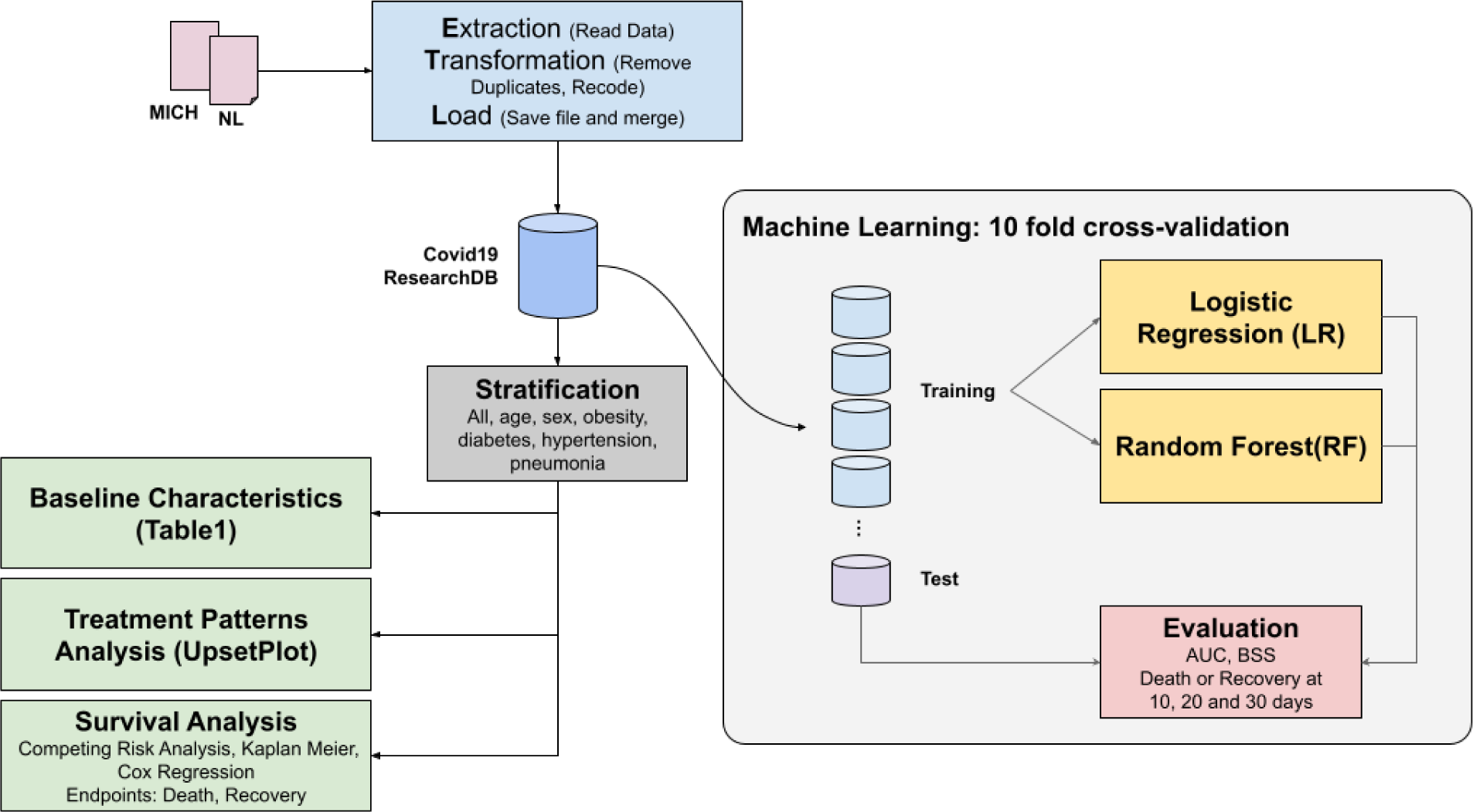
Study design. The ingestion of COVID-19 data is shown in blue, the survival analysis is shown in green, and the machine learning modeling is shown in yellow.

### Data Sources

SINOLAVE was originally developed for the influenza pandemic of 2009, and it has been quickly evolving to include new variables that are specific to COVID-19, while keeping all those that are also relevant for influenza. De-identification was done by co-author SGG, using the linkage method. Name, last names, and Social Security Number (SSN) were used. All other co-authors did not have access to identifiable information. We filtered the data to select the COVID-19 patients only, which were diagnosed either by a PCR test or by rapid antigen testing.

The database includes information about comorbidities, initial symptoms and drugs taken, also the dates of start of the clinical picture, admission, and discharge. Medication prescription is recorded in SINOLAVE in three categories: antibiotics, antivirals, and antipyretics. We used the first two categories only, since almost all patients were taking some form of antipyretic, most frequently paracetamol. The information is structured and registered at each site using structured vocabulary. All drugs and variables are reported in categories described in Supplementary Material. All missing data were considered to not be observed, and therefore assigned a missing value (no condition, symptom or drug observed).

### Statistical analysis

We calculated the cohort’s baseline characteristics for the COVID-19 positive patients, including those with antibiotic and antiviral prescriptions. We created a frequency table from the reported treatments in the database, including combinations of antibiotics and antivirals. The treatments (alone or in combination) included in the subsequent analyses were those that had at least 100 patients taking them. Time zero for these analyses was the start day of the clinical symptoms.

Building upon the guidance provided by McCaw et al. (*18*), we conducted a competing risk analysis. This approach was chosen as the classical Kaplan-Meier analysis and Cox proportional hazard model may not be suitable when confronting two competing outcomes. Given that patients who succumb to the illness cannot subsequently recover, the study inherently involves a competing risk scenario. To navigate this, we employed the cumulative incidence function to better account for and analyze the complex interplay of distinct outcomes in our investigation (*19*, *20*).

We used the CIF approach to compare the two outcomes of death and recovery stratifying by each type of treatment in the database which was taken by 100 patients or more. The Kaplan-Meier was used in both cases, and to reduce bias in the analysis we stratified the database using six variables (age group, sex, obesity, diabetes, hypertension and pneumonia). A Cox proportional hazard model was built using the following covariates: age group, sex, hospitalization status, initial symptoms and comorbidities. The odds ratio was calculated by adjusting with the same variables. To address potential sources of bias, we stratified the dataset by the most common comorbidities (i.e. diabetes, obesity, hypertension, coronary heart disease, and pneumonia), and we also performed a time-window-based study to minimize potential time-related biases.

### Predictions of mortality and recovery

We implemented two classical machine learning models to predict mortality and recovery at fixed points in time. We used a logistic regression model and a random forest model to predict the outcome of the patient (death or recovery) at 10, 20, and 30 days after the start of the clinical picture (day zero). We evaluated these classification tasks using the area under the receiver operating characteristic (AUC) with a 95% confidence interval (CI). The brier skill score (*21*) was used to test the calibration of these models. It provides an index between −1 (not calibrated) and +1 (calibrated).

## 3. Results

The dataset consisted of 130,216 COVID-19 patients with a positive real-time polymerase chain reaction (RT-PCR) test of SARS-CoV-2 (97%) or by clinical diagnosis of COVID-19 (3%). We encountered a small number of duplicates in the data which were removed from the analysis. Table 1 summarizes the baseline characteristics, comorbidities and outcomes for these patients.

**Table 1.** Baseline characteristics. Sex, Age, Comorbidities, Complications, and Mortality are shown for the main cohort and subgroups.

|  | All Covid+<br>patients | Patients taking<br>antivirals AND/OR<br>antibiotics | Patients taking<br>antivirals AND<br>antibiotics |
| --- | --- | --- | --- |
| **N** | 130,216 (100%) | 5,575 (100%) | 955 (100%) |
| **Sex** |  |  |  |
| Female | 66,445 (51%) | 2,495 (45%) | 380 (40%) |
| Male | 63,771 (49%) | 3,080 (55%) | 575 (60%) |
| **Age** |  |  |  |
| < 20 | 6,654 (5%) | 114 (2%) | 5 (<1%) |
| [20, 40) | 60,611 (47%) | 1,488 (27%) | 131 (14%) |
| [40, 60) | 43,537 (33%) | 2,027 (36%) | 360 (38%) |
| > 60 | 19,414 (15%) | 1,946 (34%) | 459 (48%) |
| **Comorbidities** |  |  |  |
| Hypertension \| Obesity \| Diabetes \| Cardiovascular | 36,808 (28%) | 2,949 (53%) | 635 (66%) |
| COPD \| Asthma \| Smoking History | 10,509 (8%) | 625 (11%) | 94 (10%) |
| Renal failure \| Chronic hepatic disease | 2,464 (2%) | 300 (5%) | 50 (5%) |
| Immunosuppression \| HIV \| TB \| Cancer | 1,746 (1%) | 128 (2%) | 24 (3%) |
| Neurological | 159 (<1%) | 11 (<1%) | 2 (<1%) |
| **Complications** |  |  |  |
| Pneumonia | 5,409 (4%) | 2,128 (38%) | 429 (45%) |
| **Death events** | 11,132 (9%) | 1,724 (31%) | 461 (48%) |

Out of these patients, only 2,746 were taking an antiviral and/or an antibiotic drug. Fluoroquinolones were the largest group of antibiotics prescribed, while neuraminidase inhibitors (NAIs) were the most frequent antivirals. Figure 2 shows the odds ratio from the demographic and symptoms variables (panel A) and from the treatments (panel B), using mortality as the endpoint. We highlighted those covariates with a more significant p-value.

**Figure 2.**
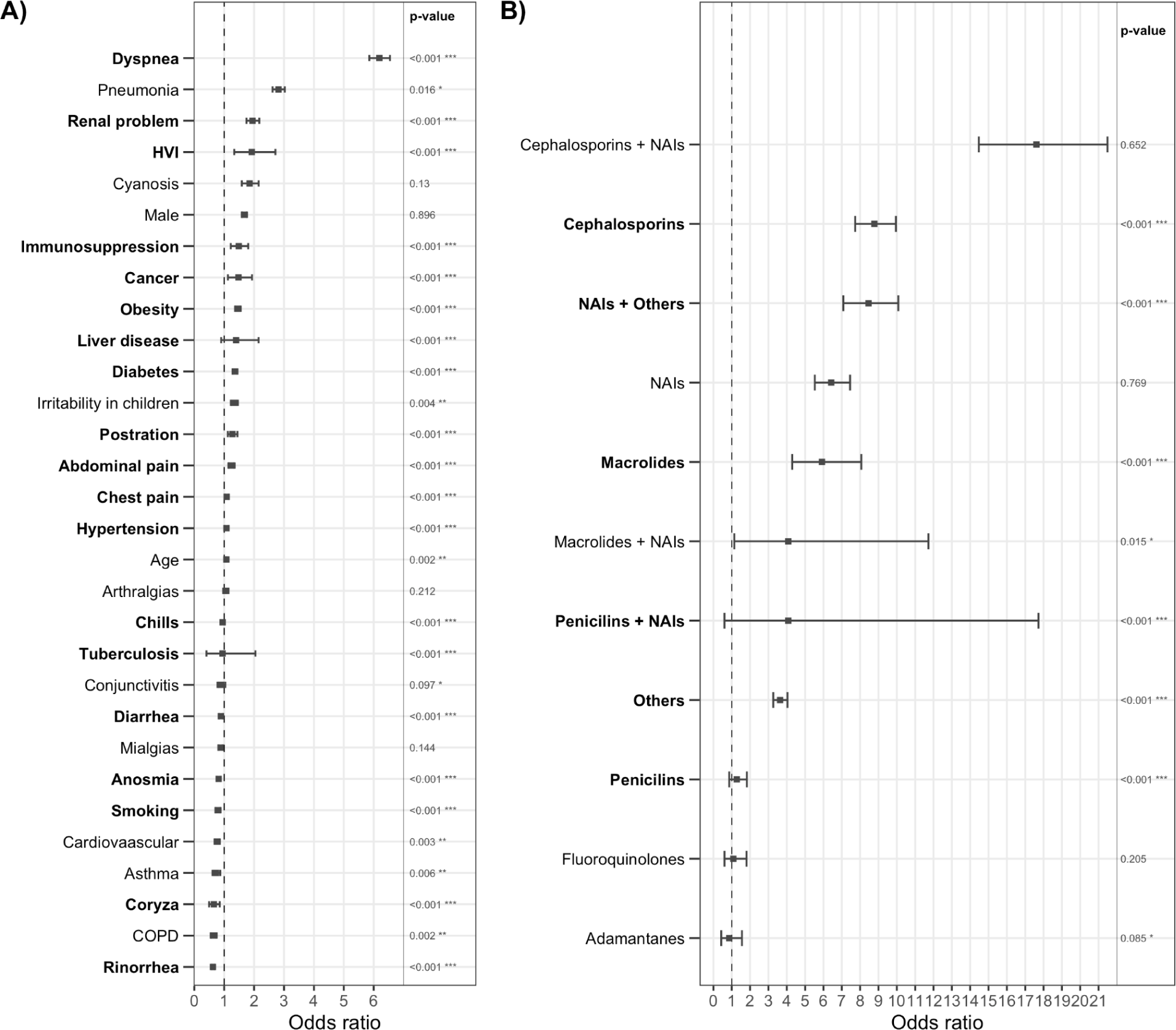
Forest plot. A) Symptoms, age and sex, B) Type of treatment. Most statistically significant variables in each method were highlighted.

The competing risk analysis depicting survival and recovery is illustrated in Figures 3. In Figures 4-5, an analysis employing Kaplan-Meier curves, stratified by various cofactors such as hypertension, age groups, diabetes, and sex, consistently yielded identical conclusions. Notably, our findings revealed that the concurrent use of neuraminidase inhibitors (NAIs) with Cephalosporins was associated with significantly lower rates of both survival and recovery compared to alternative treatment patterns. Conversely, the administration of penicillin exhibited comparable or superior rates of survival and recovery compared to patients not receiving any antibiotic or antiviral interventions. These robust findings underscore the potential impact of specific treatment combinations on patient outcomes in the context of COVID-19.

**Figure 3.**
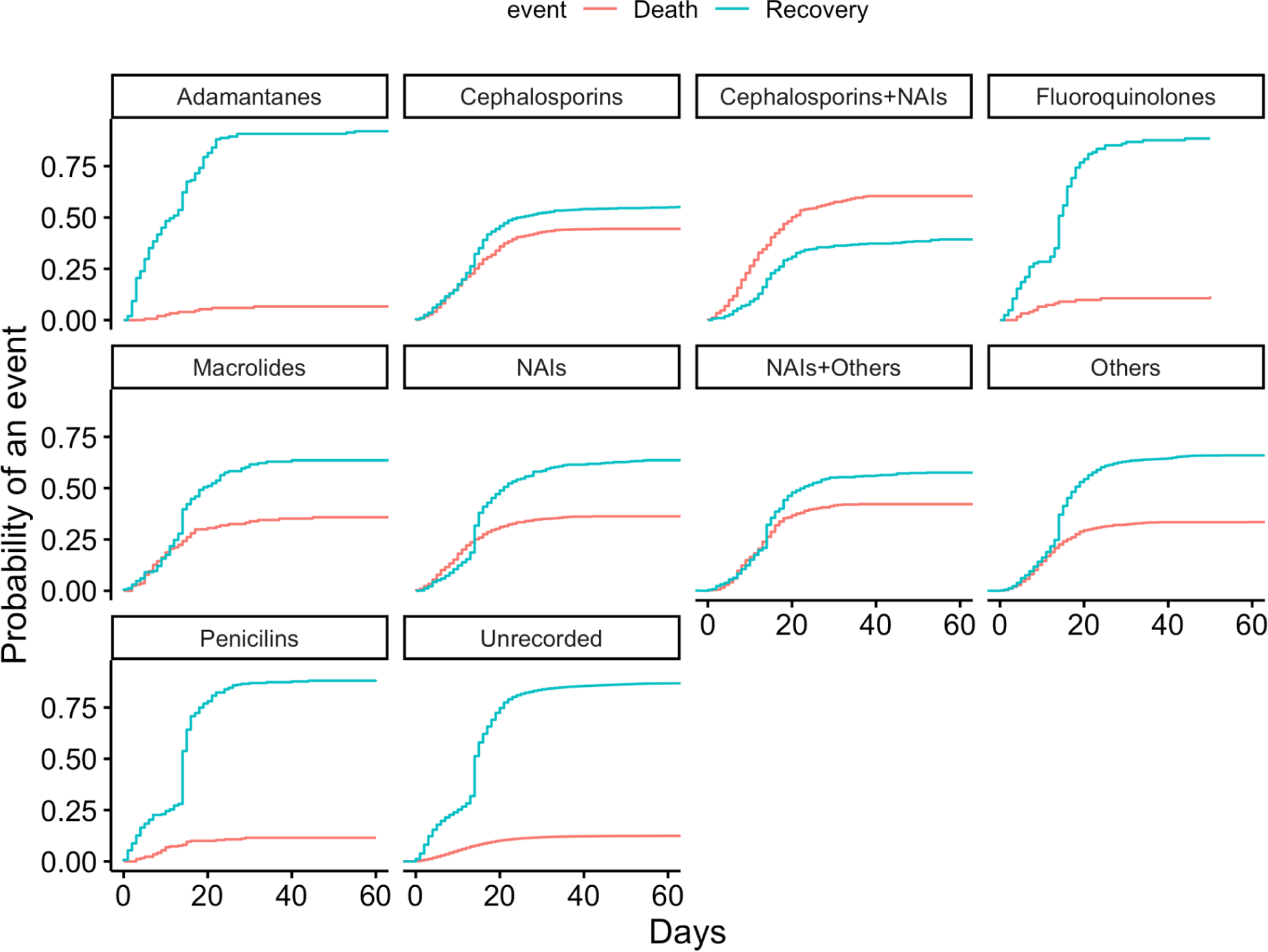
Competing Risk Analysis. Cumulative Incidences Functions for Mortality and Recovery across the different combinations of treatments. Notice that the only time that the curve of mortality surpasses the curve of recovery is in the case of the combination of cephalosporins with NAIs.

**Figure 4.**
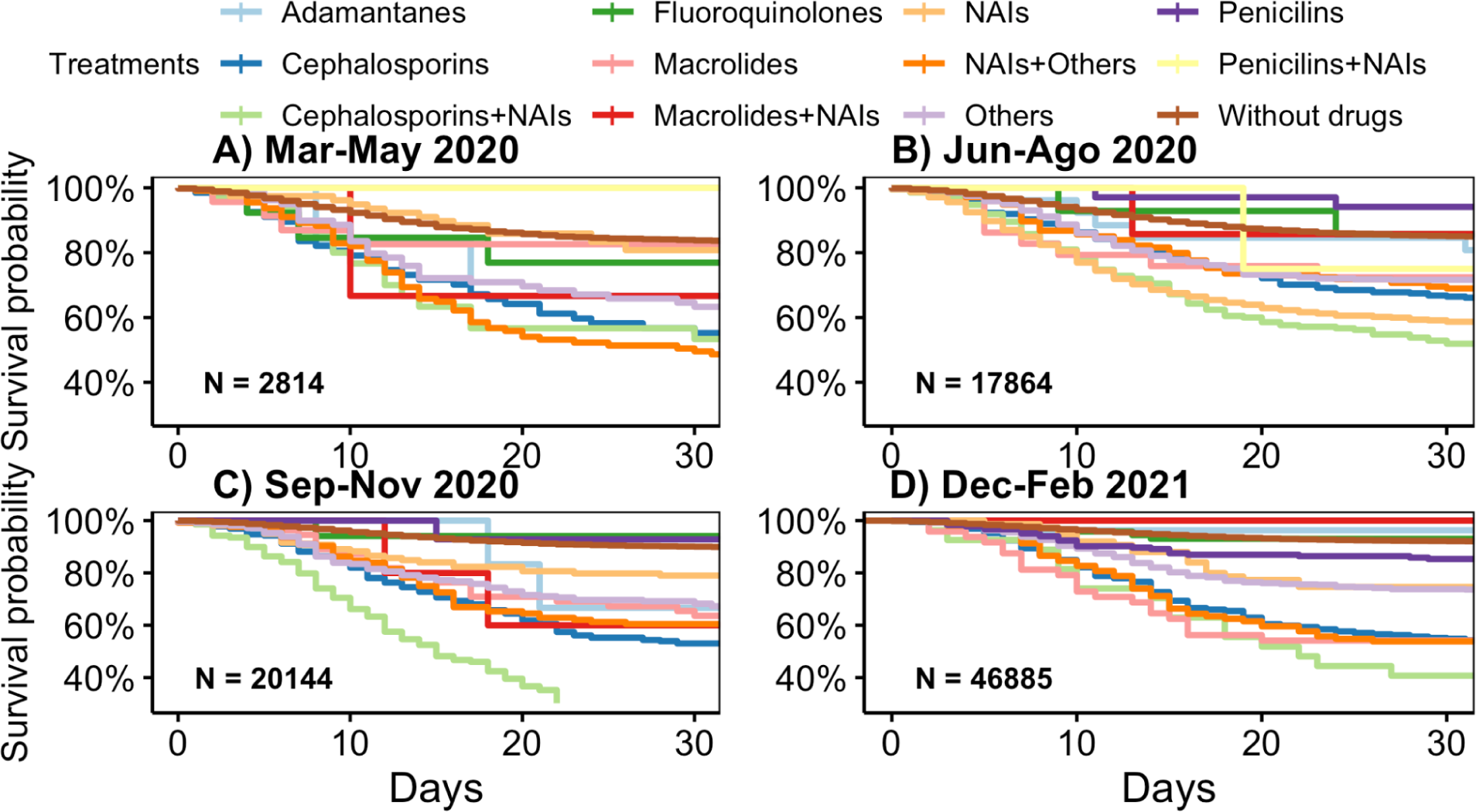
Survival analysis: 3-month time window.

**Figure 5.**
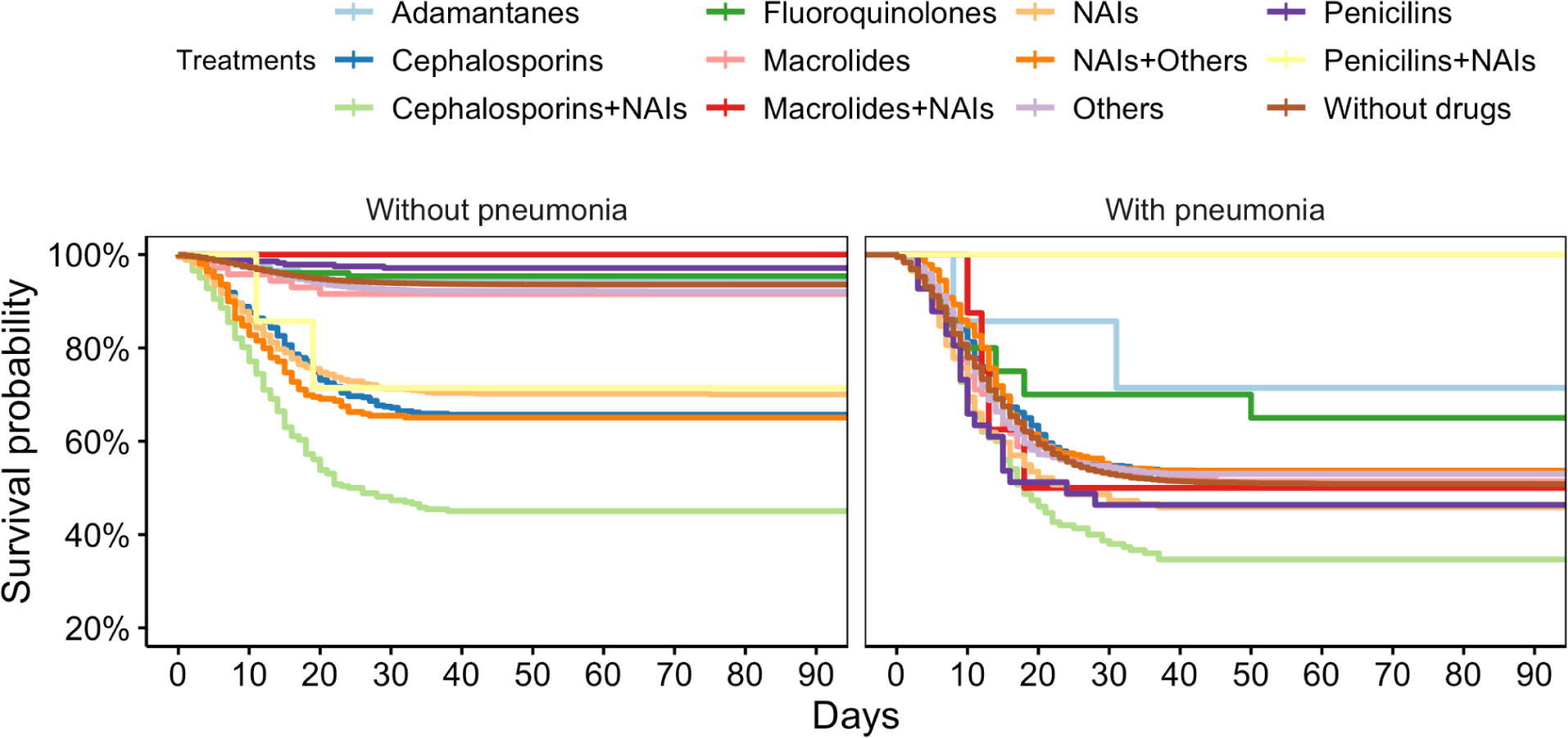
The effect of pneumonia in the survival rate of the population is clear

Contrasting with logistic regression (LR) having an average AUC of 0.85 versus the random forest (RF) with an average AUC of 0.92 the random forest model showed good performance when predicting mortality as it is shown in Table 2. The prediction accuracy improved while we relaxed the time window used (i.e., 10, 20, 30 days). All mortality models were calibrated (BSS larger than 0). In contrast, our models had a lower prediction power when predicting recovery in the same cohort, with LR achieving an average AUC of 0.69, and RF with an average AUC of 0.79. The variables used for this prediction task consisted of the initial symptoms, comorbidities and type of treatment. Lower prediction scores in the case of recovery can be explained by the fact that these patients tend to be censored more frequently. So the prediction of this becomes a harder task.

**Table 2.** AUROC curves. The area under the receiver operating characteristic (AUROC), the 95% confidence intervals (C.I.) and the Brier Skill Score (BSS) for each case is shown.

| **Classification task** | **Logistic regression** |  |  | **Random forest** |  |  |
| --- | --- | --- | --- | --- | --- | --- |
| **Mortality** | **AUC** | **95% CI** | **BSS** | **AUC** | **95% CI** | **BSS** |
| Death within 10 days | 0.83 | 0.83-0.84 | 0.09 | 0.91 | 0.91-0.92 | 0.30 |
| Death within 20 days | 0.85 | 0.85-0.85 | 0.20 | 0.91 | 0.91-0.92 | 0.38 |
| Death within 30 days | 0.86 | 0.85-0.86 | 0.23 | 0.92 | 0.91-0.92 | 0.40 |
| **Recovery** |  |  |  |  |  |  |
| Recovered within 10 days | 0.57 | 0.57-0.58 | -0.01 | 0.77 | 0.77-0.78 | 0.15 |
| Recovered within 20 days | 0.66 | 0.65-0.66 | 0.08 | 0.77 | 0.76-0.77 | 0.21 |
| Recovered within 30 days | 0.69 | 0.68-0.69 | 0.10 | 0.79 | 0.79-0.79 | 0.24 |

## 4. Discussion

In this study, in patients with COVID-19, we found a direct association of the severity of the disease with chronic disorders such as diabetes, hypertension, obesity, neoplasms, autoimmune diseases and CKD; the signs and symptoms that best predict the disease were identified, also it was found that patients who received adamantane, fluoroquinolone or penicillin as a treatment for SARS-COV-2 infection presented better survival probability, while those who received cephalosporins alone or associated with neuraminidase inhibitors show the worst survival rates.

The COVID-19 infection very rapidly reached pandemic conditions (*22*) during 2021 and continues to the present time. Several studies have associated chronic diseases such as obesity, diabetes, hypertension, and CKD, among others (*23–25*) with the development of severe forms of COVID-19 infection, including pneumonia. Mexico ranks third worldwide in deaths attributed to COVID-19 infection and with a high prevalence of these diseases, including CKD (*26*); therefore, strict control of these conditions is necessary in patients with COVID-19 infection in Mexico and the world.

Regarding access to medicines in the IMSS and treatment policies available to IMSS doctors, these evolved rapidly during the pandemic emergency. As mentioned in the Introduction section, the guidance offered by the Ministry of Health (MoH) was quickly adapted to the global knowledge available at the time. Symptomatic patients with cough, fever, dyspnea or headache were promptly followed up for further investigation including PCR or antigen testing and other laboratory tests. However, due to the limited resources available in a public health system, access to investigational medicinal products from clinical trials was limited. Also, it was unclear at the time what effect any drug or treatment would have had on this new virus. However, the beneficial effect observed in the survival rate in patients who received adamantane, fluoroquinolone, or penicillin is striking. Special mention is the development of vaccines. Although the vaccination program officially started in January 2021 in Mexico with health workers, the effects of this vaccination program were not observed in our data (March 2021 cutoff).

Regarding the relevant cofactors affecting survival in the present study, we observed that medication patterns differed when different scenarios (e.g., diabetes, hypertension, age, gender) were considered, even when the treatments studied had no known contraindications for the subgroups of the population investigated. The impact of medication on survival outcomes is complex in a real-life setting. Various cofactors could influence it, and various reasons could explain these findings. First, the impact of preexisting comorbidities on the response to infection and target treatment should be considered. Age and age-related diseases, including type 2 diabetes, obesity, and cardiometabolic diseases, are well-known risk factors for the severity of COVID-19 (*27–29*). These conditions are associated with a chronic inflammatory state (*30*) and reduced innate and adaptive immune responses (*31*, *32*). In fact, these population subgroups have also been associated with decreased influenza virus survival (*33*). Thus, the patient’s medical history, as well as the intrinsic individual variability in the response to medication, or the pharmacogenomic profile, are relevant cofactors that influence individual medication patterns. Furthermore, patients with multiple comorbid conditions may be taking multiple medications, that is, they may have higher rates of polypharmacy. The potential effect of drug-drug interactions (DDIs), how DDIs present clinically, and their direct or indirect effect on COVID-19 outcomes should be considered. Although the underlying biological mechanism triggering these differences in the context of COVID-19 remains largely unknown, in the near future, the integration of these different layers of information should promote better clinical management of COVID-19 patients. On the other hand, the Mexican public health authorities recently updated the clinical guideline for the treatment of COVID-19, including new antiviral drugs approved by the U.S. Food and Drug Administration (FDA) at the end of 2021 (19,20). These drugs were molnupiravir for adults with a positive test result and with a high risk of developing severe forms of the disease, and paxlovid for the treatment of mild-to-moderate cases in adults and pediatric patients. Moreover, the widespread adoption of these new antivirals is not entirely clear despite being oral drugs for outpatient use, as they are more accessible than drugs administered intravenously (remdesivir, sotrovimab), which require hospital monitoring or are high-cost drugs (*36*).

### On the electronic healthcare system

This study was only possible due to the digitization of SINOLAVE, an IMSS database that was developed during the 2009 Influenza pandemic and later on refurbished to serve the information needs of the 2019 COVID-19 pandemic. Furthermore, the emergence of virtual randomized clinical trials has shown the value to accelerate results and provide informed guidance to patients (*37*). The digitization of electronic health records is possible and should be an overall strategy in resource-constrained settings to provide a data-driven approach to public health concerns (*38*). Machine learning models and data science can be effectively applied to these settings augmenting the capabilities of the healthcare system.

### On the prediction of mortality and recovery

The positive score while predicting mortality suggests that the variables used by the model explain with satisfactory confidence the outcome of the patient at 30 days from the onset of symptoms. On the other hand, the recovery prediction was not as good, this suggests that the time to recovery could be driven by other factors.

### Limitations

This study has several limitations. One limitation is having obtained the information from the SINOLAVE registry. As previously mentioned, the SINOLAVE record was originally structured to generate information on other viral diseases, so it has been adapted to generate information related to COVID-19, including several pharmacological treatments, for the specific purpose of studying the effect of COVID-19 treatments; therefore, other drugs not listed in the registry cannot be analyzed and can be possible confounding factors. Also, the chaotic environment in the pandemic could have affected the quality ingestion of the data in the registry, many rushes were made during the early stages of the pandemic. Another limitation is the short study period used in our study, so the late post-COVID clinical manifestations are unknown. A third limitation is that the study period was set before the vaccination of the general population, so the current treatment conditions may be different.

## Conclusion

We observed that the use of the combination of NAIs and cephalosporins as a treatment for COVID-19 presented the lowest survival probability by a considerable margin. It is possible that this effect is observed partly due to a more susceptible population within this group (e.g. higher number of comorbidities), or that the treatment was prescribed after the progression of the disease. Still, we would not recommend the use of this specific combination since it showed the worst survival rate by a great margin, even in those patients without comorbidities, as well as in the younger population. All the other combinations showed much more beneficial outcomes. Surprisingly the use of penicillin alone showed one of the best survival chances for COVID-19 patients in our study, we have not a straightforward explanation for this fact other than the possible prevention of pneumonia which was one of the main complications leading to death.

## Supporting information

Supplemental Table 1. Variables

## Acknowledgements

Authors would like to thank Dr. Noga Or-Geva, Dr. Sonia Moreno-Grau, and Dr. Carlos D. Bustamante for their valuable insights when designing this manuscript.

## Contributions

ALP and CAA designed the study. SGG and GGA did the data curation. GGA, CAA, AGG, SGG, and ALP performed the analysis. GGA, AGG, SMG, CAA, and ALP drafted the manuscript. All authors contributed critically, read, revised and approved the final version.

## Ethics approval

This study was approved by the Ethics and Research Committees from the Mexican Institute of Social Security (IMSS), which are certified as Institutional Review Board (IRB) in accordance with the Mexican regulation, under protocol numbers R-2021-1912-014, R-2020-785-058. Since, this was a retrospective study and the author SGG carried out the anonymization of data, the IRB waived all need for consent for this study.

## Data Availability

The data that supports the findings of this study is available for research purposes upon written request, which will be reviewed on a case-by-case basis by IMSS IRB given the submission of a research protocol.

